# Longitudinal immune profiling after radiation-attenuated sporozoite vaccination reveals coordinated immune processes correlated with malaria protection

**DOI:** 10.1101/2022.09.12.22279869

**Authors:** Fergal J Duffy, Nina Hertoghs, Ying Du, Maxwell L. Neal, Damian Oyong, Suzanne McDermott, Nana Minkah, Jason Carnes, Katharine V Schwedhelm, M Juliana McElrath, Stephen C De Rosa, Evan Newell, John D. Aitchison, Ken Stuart

## Abstract

Identifying immune processes required for liver-stage sterilizing immunity to malaria infection remains an open problem. The IMRAS trial comprised 5x immunizations with radiation-attenuated sporozoites resulting in 55% protection from subsequent challenge. To identify correlates of vaccination and protection, we performed detailed systems immunology longitudinal profiling of the entire trial time course including whole blood transcriptomics, detailed PBMC cell phenotyping and serum antigen array profiling of 11 IMRAS RAS vaccinees at up to 21 timepoints each. RAS vaccination induced serum antibody responses to CSP, TRAP, and AMA1 in all vaccinees. We observed large numbers of differentially expressed genes associated with vaccination response and protection, with distinctly differing transcriptome responses elicited after each immunization. These included inflammatory and proliferative responses, as well as increased abundance of monocyte and DC subsets after each immunization. Increases Vδ2 γδ T cells and MAIT cells were observed in response to immunization over the course of study. Interferon responses strongly differed between protected and non-protected individuals with high interferon responses after the 1^st^immunization, but not the 2^nd^-5^th^. Blood transcriptional interferon responses were correlated with abundances of different circulating classical and non-classical monocyte populations.

This study has revealed multiple coordinated immunological processes induced by vaccination and associated with protection. Our work represents the most detailed immunological profiling of a RAS vaccine trial performed to date and will guide the design and interpretation of future malaria vaccine trials.

## INTRODUCTION

Currently, there exists a single approved malaria vaccine: the subunit-based RTS’S/AS01 (aka Mosquirix) [1]. The RTS’S antigen consists of portions of the circumsporozoite protein (CSP) fused to the viral surface antigen of hepatitis B. However, the efficacy of this vaccine is low: reported as 35.9% i(95% CI, 8.1% to 55.3%) in the first year after vaccination for young children [2], and further reduced at later times. Given that the burden of malaria deaths falls most severely on young children in endemic areas, an improved vaccine is urgently needed. Recently, the R21/MM vaccine candidate, also a CSP protein-based vaccine, has shown promise in this area exhibiting 74-77% efficacy [3], however, the study follow-up period was short and the number of individuals enrolled was relatively small.

A key advantage of CSP as a vaccine antigen is that it is highly expressed on the *Plasmodium falciparum* sporozoite, the form of the parasite injected into the skin by mosquito bite that goes on to infect the liver. The sporozoite represents a population bottleneck, as only low numbers of sporozoites successfully infect the liver and go on to induce a symptomatic blood stage infection. Hence, a successful liver-stage specific vaccine would prevent symptomatic disease and transmission and allow the immune system to face a relatively small number of parasites. Whole-sporozoite vaccines represent an alternative approach to CSP-subunit vaccines. Whole-sporozoite vaccines require the administration of entire sporozoites, which are difficult to mass produce compared to a more traditional protein subunit vaccine platform. However, whole sporozoite vaccination presents an enhanced antigenic repertoire while still targeting the low-parasite burden liver stage. It has long been observed that radiation-attenuated sporozoites are capable of inducing sterilizing immunity against live challenge in both animal and human malaria infection [4–7]. Along with radiation attenuation, sporozoite administration under anti-parasite chemoprophylaxis [8,9] and genetic attenuation [10,11] have also been shown to be effective at preventing infection. Notably, sporozoite administration under chemoprophylaxis has been shown to be capable of eliciting strain-transcendent protection against subsequent challenge [12].

Previous work has described the IMRAS study [13–15], which comprised radiation-attenuated sporozoite vaccination of malaria-naïve individuals who were predominantly male (10 male, 1 female). Studying malaria-naïve rather than malaria experienced individuals allows the clearest picture of the early events involved in protective immunity. It has long been observed that, unlike RAS vaccination, a blood-stage malaria infection will not result in sterilizing immunity to reinfection. but rather a slow development of malaria tolerance after repeated infections [16– 19]. Blood stage infection is associated with impaired development of immunity to pre-erythrocytic malaria [20,21].

The IMRAS study was designed with the goal of delivering an intermediate dose of sporozoites that would induce protection in roughly half of the vaccinees, resulting in 55% protection in our study cohort. We have previously published our systems biology analysis of responses to the initial priming immunization in this cohort [13]. The most striking protection-associated difference we observed was a sharp increase in type I interferon signaling 1 day after immunization in non-protected individuals. Here, we extend our analysis to a comprehensive set of timepoints after all 5 immunizations and incorporate serum antibody profiling in an attempt to disentangle the complex and highly variable blood transcriptional and cell type abundance dynamics that are associated with RAS-induced protection. We saw that trancriptome responses to RAS differed strongly after each immunization. Populations of multiple cell types including Vδ2 γδT cells and MAIT cells expanded over the course of immunizations. Furthermore, the association of increased interferon with non-protection was specific to immunization 1, with immunizations 2-5 showing a positive correlation with interferon associated gene transcription and protection. Interferon’s association with protection was correlated with different classical and non-classical monocyte abundances after each vaccination.

## METHODS AND MATERIALS

### Ethics statement

The study was conducted at the Naval Medical Research Center (NMRC) Clinical Trials Center from 2014 to 2016; controlled human malaria infections (CHMIs) were conducted at the Walter Reed Army Institute of Research (WRAIR) secure insectary. The study protocol was reviewed and approved by the NMRC Institutional Review Board in compliance with all federal regulations governing the protection of human subjects. WRAIR holds a Federal-wide Assurance from the Office of Human Research Protections (OHRP) under the Department of Health and Human Services as does NMRC. NMRC also holds a Department of Defense/Department of the Navy Federal-wide Assurance for human subject protections. All key personnel were certified as having completed mandatory human subjects’ protection curricula and training under the direction of the WRAIR Institutional Review Board or the NMRC Office of Research Administration (ORA) and Human Subjects Protections Branch (HSPB). All potential study subjects provided written, informed consent before screening and enrollment and had to pass an assessment of understanding. This study was conducted according to the Declaration of Helsinki as well as principles of Good Clinical Practices under the United States Food and Drug Administration Investigational New Drug (IND) application BB-15767. This trial was performed under an IND allowance by the Food and Drug Administration (FDA).

### IMRAS study design

The IMRAS trial (ClinicalTrials.gov NCT01994525) design has been described previously [13–15]. In this work, we analyzed IMRAS Cohort 1, which consisted of 11 individuals who received 5 RAS immunizations via approximately 200 mosquito bites each. Protective efficacy was determined by CHMI with bites from 5 NF54 *P. falciparum* infected mosquitos. Protected individuals were defined as those that remained free of detectable blood stage malaria infection via PCR and thick-blood smear post-CHMI. These vaccinees were accompanied by 3 mock immunized individuals (mosquito bites only) who served as infectivity controls, all of whom were not protected after CHMI.

### Serum antibody profiling via *P. falciparum* antigen protein array

Serum samples were taken from the 11 IMRAS RAS vaccinees and 3 mock-immunized study participants at day 0 (pre-vaccination) and day 140 (3 weeks after the fifth immunization and immediately pre-CHMI). Serum was profiled on *P. falciparum* whole-proteome microarrays, (PfWPM: Antigen Discovery Inc, Irvine CA), consisting of 7,455 *P. falciparum* full or partial proteins, representing 4,805 unique genes (91% of the *P. falciparum* proteome), to quantify levels of *P. falciparum* antigen-specific serum antibodies. *P. falciparum* array antigens were produced using either a cell-free ORF-expression clone library followed by *in vitro* transcription/translation (IVTT). After profiling, per-spot microarray signal intensity (MFI) was normalized by taking median foreground intensity minus median background fluorescence intensity based on IVTT control spots subtracted, and log2 transformed. To identify vaccine-induced serum antibody responses, the normalized MFI response was calculated as the increase in normalized MFI per-participant from day 0 to day 140 for RAS-immunized individuals. Significantly induced antibody responses were identified as those increased at least 2-fold from day 0 to day 140 with an FDR-adjusted single tailed t-test significance threshold of 0.05.

### Whole blood RNAseq

Whole blood RNAseq was performed as previously described [13] on 11 IMRAS Cohort 1 RAS vaccinees. Whole blood was collected directly into PAXgene blood RNA tubes (PreAnalytiX, Hombrechtikon, Switzerland) and stored at −20°C. RNA extraction and globin transcript depletion (GlobinClear, ThermoFisher Scientific, MA, USA) were performed prior to cDNA library preparation using the Illumina TruSeq Stranded mRNA sample preparation kit (Illumina, CA, USA). Globin transcript depletion, cDNA library preparation and RNA sequencing were performed by the Beijing Genomics Institute (Shenzhen, China). A total of sixty-six RNAseq samples were sequenced, with a target depth of 30 million reads per sample. Samples from four timepoints (day 0, 35, 119, 140) were sequenced on Illumina (San Diego, CA) Hiseq2000 sequencers using 75 base-pair (bp) paired-end reads. The remaining samples were sequenced on BGI500 sequencers using 100 bp paired-end reads.

### RNAseq differential expression and gene-set enrichment analysis

Raw RNAseq fastq files were filtered for quality control by adjusting base calls with phred scores < 5 to ‘N’, and read pairs for which either end had fewer than 30 unambiguous base calls were removed. Read pairs were aligned to the human genome (hg19) using STAR (v2.3.1d) [22]. Gene count tables were generated using htseq (v. 0.6.0) [23] with the intersection-strict setting on and Ensembl gene annotations (GRCh37.74) used to link genomic locations to gene identifiers. Raw aligned RNAseq counts were normalized to account for different sequencing platforms (Hiseq2000 vs BGI500) using ComBat-seq [24], taking advantage of 4 samples resequenced on both sequencers to ensure an unbiased data transformation. Principal component analysis was performed on log-transformed counts-per-million gene expression data.

The R limma and voom packages [25,26] were used to identify differentially expressed genes augmented by the dream package [27] to allow the use of a mixed modelling approach. Timepoint and protection were set as fixed effects, including interaction terms, with a random intercept for each participant. For each gene, models were fit using the formula:

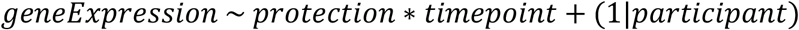

To identify genes significantly changed in expression relative to the day of most recent immunization, i.e. vaccine-induced genes, contrasts were constructed of the form ‘timepoint – most recent immunization timepoint’, e.g. ‘D003 – D000’ represents changes at day 3 relative to day 0 (day of first immunization). An FDR cutoff of 0.05 was used to identify significant genes at all timepoints. Protection-associated genes were identified by F-test, as implemented in the limma topTable function, to identify genes that where any combination of protection:timepoint interaction coefficients were significant.

Gene-set enrichment analysis (GSEA) was performed using the R fgsea package [28], using the BloodGen3 coherently expressed blood transcriptional modules (BTMs) [29]. Gene rankings used as input for GSEA were based on estimated log2 fold changes obtained from the limma-dream model. The limma topTable function was used to extract fold changes for each ‘timepoint – most recent immunization induced timepoint’ contrast, which were input to fgsea to calculate vaccine-induced BTM enrichment. Similarly each protection:timepoint interaction coefficient was used to input fold changes into fgsea to to identify enriched protection-associated BTMs.

BTM expression was calculated as the 25% trimmed mean of log-transformed counts-per-million gene expression of each BloodGen3 BTM gene, per sample. BTMs with reliable expression in less than half of their genes were excluded. Non-coherent BTMs, defined as BTMs where less than half of their genes were strongly correlated (Pearson’s r > 0.5) with the trimmed mean of BTM expression were also excluded.

### PBMC flow cytometry profiling

Flow cytometry profiling was carried out using previously published antibody panels and gating strategies [13,30,31]. PBMCs were collected from IMRAS participants and frozen for later use. After thawing in RPMI supplemented with 10% FBS and benzonase nuclease (Millipore EMD 0.05 U/ml), the samples were incubated with LIVE/DEAD Fixable Blue Dead Cell Stain Kit and the Human BD Fc Block for 30 min at room temperature before being simultaneously stained with four phenotyping panels as shown in OMIP-044 [30], OMIP-064 [31] and **Table S 1**. The cells were then profiled using a BD FACSymphony flow cytometer. The data were analyzed, and cellular populations gated and quantified as previously using FlowJo Software (version 9.6.6). The abundance of each manually gated cell subset was calculated as a percentage, using the counts of each defined cell subset divided by the total single live cells from that sample.

### Statistical analysis of flow-cytometry cell counts

Mixed models were used to identify vaccine-induced (i.e. significantly changed in abundance over time) and protection-associated cell type subsets using the R lme4 package [32]. A nested approach was taken where a ‘full’ model including protection and timepoint as fixed effects, as well as interaction terms, was compared to protection-only and timepoint-only fixed effect models. In all cases, per-participant intercepts were fit as random effects.

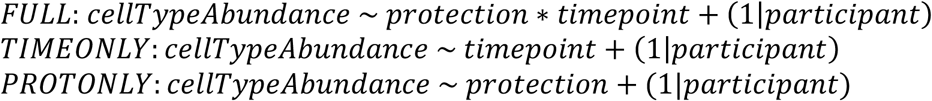

ANOVA was used to compare the FULL model to the TIMEONLY model (to determine the effect of protection on abundance) and the PROTONLY model (to determine the effect of sampling timepoint on abundance) and calculate associated p-values, and an FDR threshold of 0.05 was applied to determine significance.

## RESULTS

### The IMRAS study

The IMRAS study design has previously been described [13–15]. Briefly, study participants were immunized five times with radiation attenuated *P. falciparum* NF54 sporozoites (RAS) delivered by mosquito bite, with roughly 200 bites per-immunization. Immunizations were spaced 4-5 weeks apart with protection determined by controlled human malaria infection (CHMI) three weeks after the final immunization. In this study, we analyzed serum antibody levels, whole blood transcriptomics and PBMC cell phenotypic profiles from the 11 RAS-immunized (6 protected, 5 non-protected) participants in IMRAS Cohort 1. The cohort also included three mock-immunized individuals who received sporozoite-free mosquito bites followed by CHMI on the same schedule as RAS vaccinees. Serum, PBMCs and whole-blood PAXgene samples were obtained for up to 5 timepoints per-immunization per-participant for analysis (**Table 1**)

**Table 1:** IMRAS sampling schedule. X indicates where specific sample types were taken.

|  | Imm 1 |  |  |  |  | Imm 2 |  |  |  | Imm 3 |  |  |  | Imm 4 |  |  | Imm 5 |  |  |  | CHMI |
| --- | --- | --- | --- | --- | --- | --- | --- | --- | --- | --- | --- | --- | --- | --- | --- | --- | --- | --- | --- | --- | --- |
| Day | 0 | 1 | 3 | 7 | 14 | 28 | 31 | 35 | 42 | 56 | 59 | 63 | 70 | 84 | 87 | 91 | 119 | 120 | 122 | 126 | 140 |
| Whole blood | X | X | X | X | X | X | X | X | X | X | X | X | X | X | X | X | X | X | X | X | X |
| PBMC | X |  | X | X | X |  |  |  |  | X | X | X | X |  |  |  | X |  | X | X |  |
| Serum | X |  |  |  |  |  |  |  |  |  |  |  |  |  |  |  |  |  |  |  | X |

### RAS immunization induced detectable serum antibody responses

Serum samples were taken from all 11 vaccinees and 3 mock-immunized individuals immediately prior to immunization (day 0) and challenge (day 140). Each individual serum was screened on *P. falciparum* antigen protein arrays [Antigen Discovery Inc.] to detect the presence of *P. falciparum*-specific antibodies. Strongly significant antibody responses to three *P. falciparum* proteins: circumsporozoite protein (CSP), Apical membrane antigen 1 (AMA1) and thrombospondin related adhesion protein (TRAP) were seen in RAS immunized individuals **(Figure 1 A)**. All three of these proteins are known to be expressed on the surface of on the apical membrane of *P*.*falciparum* sporozoites, and are involved in hepatocyte invasion and binding [33–35]. Antibody responses did not significantly differ between protected and non-protected individuals (**Figure 1 B**).

**Figure 1:**
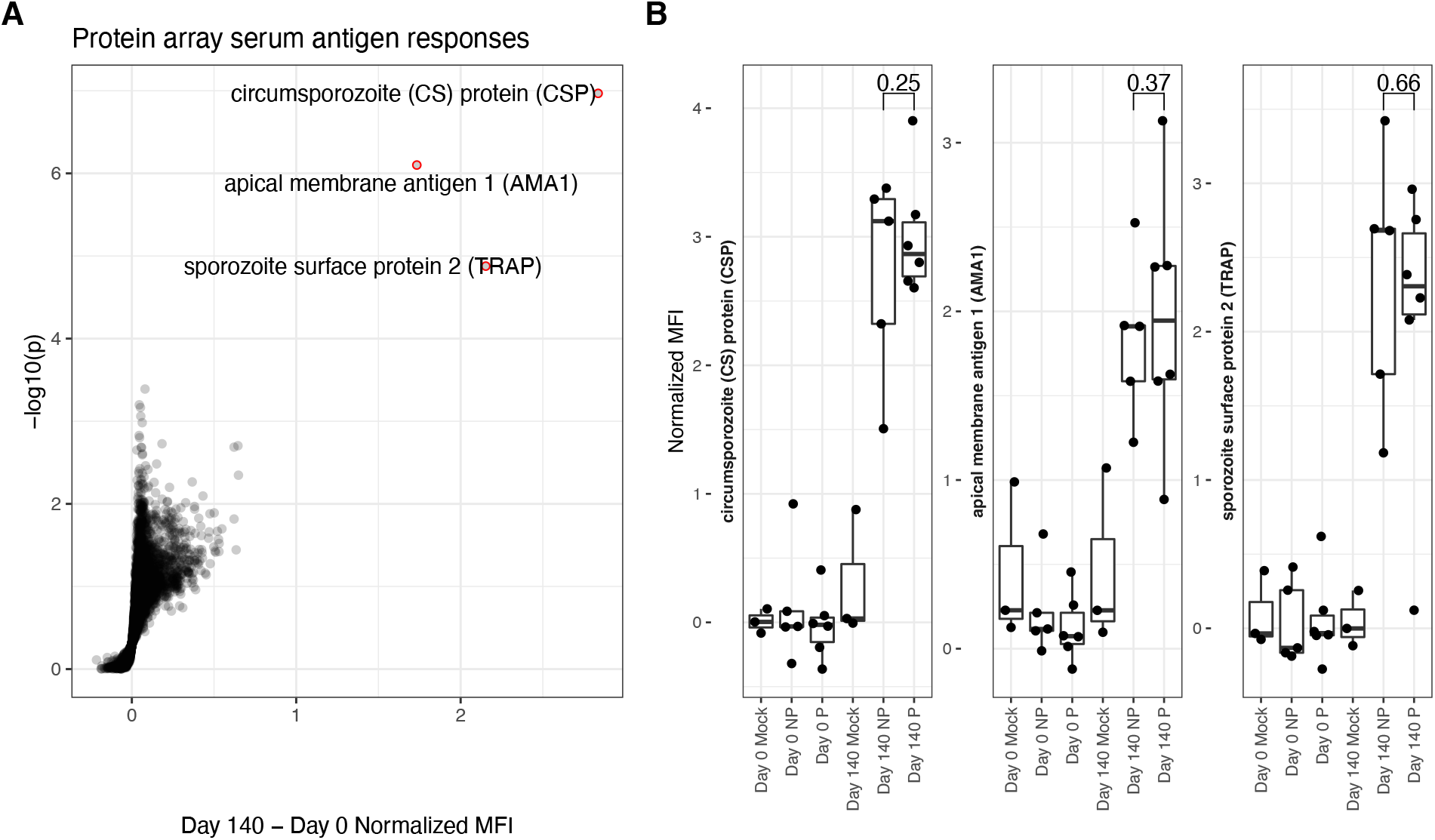
Protein array determined serum antigen responses. **A**. Volcano plot. X-axis shows increase in protein array normalized mean fluorescent intensity (MFI) from pre-immunization (day 0) to immediately pre-challenge (day 140) timepoints for RAS-immunized participants. Y-axis shows matching log-transformed false-discovery rate (FDR) adjusted single-tailed t-test p-values. Significant (FDR < 0.05) and highly increased (MFI increase > 1) antibody responses are highlighted with red circles and labelled with the corresponding protein. **B**. Box and dot-plots of pre-vaccine and pre-challenge MFIs for each study participant for labelled antibody responses from **A**. shown separately for protected (P) and non-protected (NP) RAS vaccine recipients. Mock immunized individuals are included for reference. T-test p-values comparing pre-challenge antibody binding MFIs between P and NP individuals are indicated.

### Differential expression analysis revealed vaccine-induced and protection-associated genes

To identify vaccine-induced responses that correlate with protection, we carried out longitudinal whole blood transcriptomic profiling of IMRAS RAS vaccinees over the entire course of repeated immunizations. A mixed modelling approach was applied (see **Methods**) to identify genes that were responsive to vaccination and/or associated with protection that also accounted for gene expression levels primarily attributable to per-participant effects. Principal component analysis of transcriptome profiles showed that whole blood expression profiles clustered closely together by participant, rather than by longitudinal timepoint (**Figure S1**), underscoring the need to incorporate transcriptional per-participant effects as a factor in our differential expression analysis.

Vaccine-induced differentially expressed genes (DEGs) were identified as those significantly changed in expression in blood relative to the most recent previous immunization. Hundreds of vaccine-induced DEGs were identified (FDR < 0.05) at the majority of timepoints across the entire course of immunizations (**Figure 2 A, Datafile S 1**). Most strikingly, we observed large numbers of genes decreased in expression after the second immunization. In contrast, there were relatively few vaccine induced DEGs seen after the third immunization. We quantified the overlap of these DEGs with the recently published BloodGen3 [29] coherent blood transcriptional modules (BTM) to interpret biological processes induced post-immunization. BTMs significantly enriched in vaccine-induced genes that were uniformly increased or decreased per-timepoint are shown in **Figure 2 B** (FDR-adjusted hypergeometric p-value < 0.05). Genes increased in expression in the two weeks after the first immunization were enriched for interferon, oxidative phosphorylation and erythroid cell BTMs. Genes reduced in expression after the second immunization were enriched for protein synthesis, protein modification, gene transcription and oxidative phosphorylation BTMs at multiple timepoints. Fewer significant BTMs were identified in DEGs after the 3^rd^ – 5^th^ immunizations, although BTMs representing cell death, and inflammation were increased 3 days after the 3^rd^ immunization. Numbers and expression patterns of DEGs and accompanying enriched BTMs were highly distinct after each immunization.

**Figure 2:**
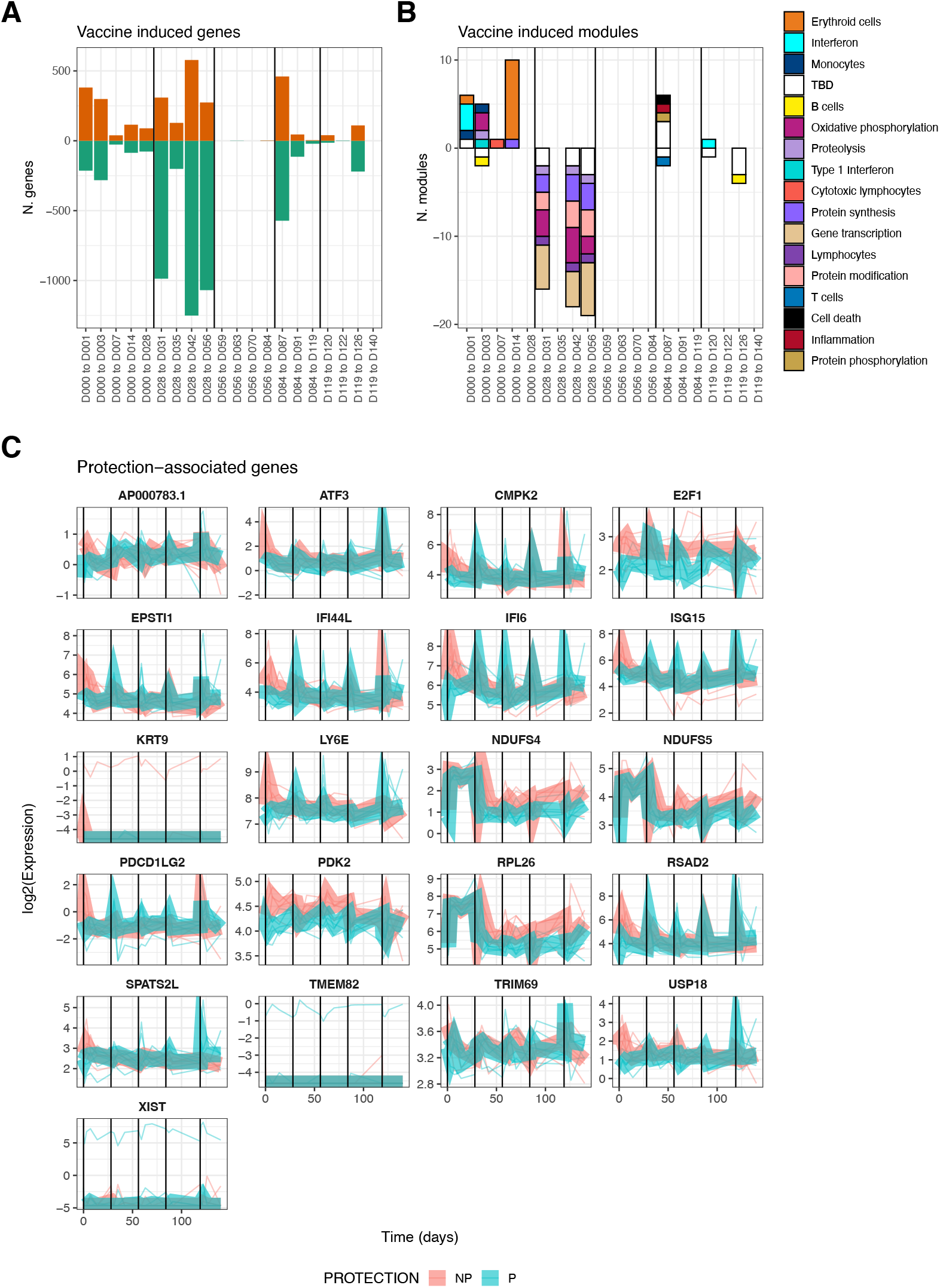
Differentially-expressed genes associated with vaccine-induced and/or protection-associated responses to immunization. **A**. Bar plot showing counts of genes significantly increased (positive) or decreased (negative bar) relative to the day of the most recent vaccination. **B**. Bar plot showing counts of BloodGen3 modules enriched in genes shown in **A**., per-timepoint and direction (positive bars represent modules enriched in genes with increased expression relative to the day of the most recent immunization, negative bars represent enrichment for genes with decreased expression). Module functional annotation is shown by the color key. **C**. Line plots showing per-individual normalized expression of each protection-associated gene over time colored by protection status. Group-averaged expression is shown as a ribbon. For all panels, vertical black lines indicate times of immunizations.

We also found 21 genes significantly associated with protection when considering all timepoints together (**Figure 2 C**). The 21 protection-associated genes significantly overlapped (FDR-adjusted hypergeometric p = 3 × 10^−8^) with a single BTM: six protection-associated genes (EPSTI1, IFI44L, IFI6, ISG15, LY6E, RSAD2) are members of BTM M8.3 representing Type I Interferon responsive genes. These genes were also increased in expression immediately post-immunization, but whereas non-protected individuals had high and sustained increased expression of these genes after the first immunization, protected individuals had stronger increases after the remaining 4 immunizations (**Figure 2 C**). This suggests that while interferon responses are broadly associated with protection during RAS immunization, the effect of increased interferon gene expression on protection may change over the course of vaccination. 13 of these 21 total protection-associated genes and 5 of 6 type I interferon protection-associated genes were also significantly vaccine-induced to for at least one timepoint.

### Gene-set enrichment analysis revealed complex timing of vaccine responses

Modules identified as having enriched overlap with DEGs were limited by the binary significance cutoff used to identify DEGs. Therefore, we applied gene-set enrichment analysis (GSEA) to identify BTMs that were significantly enriched in the full rank-ordered gene list. Genes were ranked based on the above determined linear model coefficients and normalized enrichment scores (NES) calculated to classify BTMs as being increased or decreased relative to the day of the most recent immunization (**Figure 3 A**) or associated with protection at a specific timepoint (**Figure 3 B**). Overall, vaccine-induced responses identified as enriched in DEGs (**Figure 2 B**) were also captured in the GSEA analysis; for example, increased interferon and erythroid cell BTMs after the first immunization, and decreased gene transcription and protein synthesis BTMs after the second immunization. However, the vaccine-induced GSEA captured many events after the third, fourth and fifth immunizations not seen in the DEG-based analysis. This includes increases in erythroid cell and gene transcription and decreased inflammation after the third immunization as well as increased inflammation, interferon and protein phosphorylation after the fourth and fifth immunization. Many significantly enriched BTMs fall into the ‘TBD’ category, where the overall functional annotation was not determined. These show enrichment patterns similar to inflammation and cytokine/chemokine BTMs, suggesting co-ordination with defined immune responses. Concordant with the individual DEG analysis, we saw highly distinct response patterns induced after each immunization.

**Figure 3:**
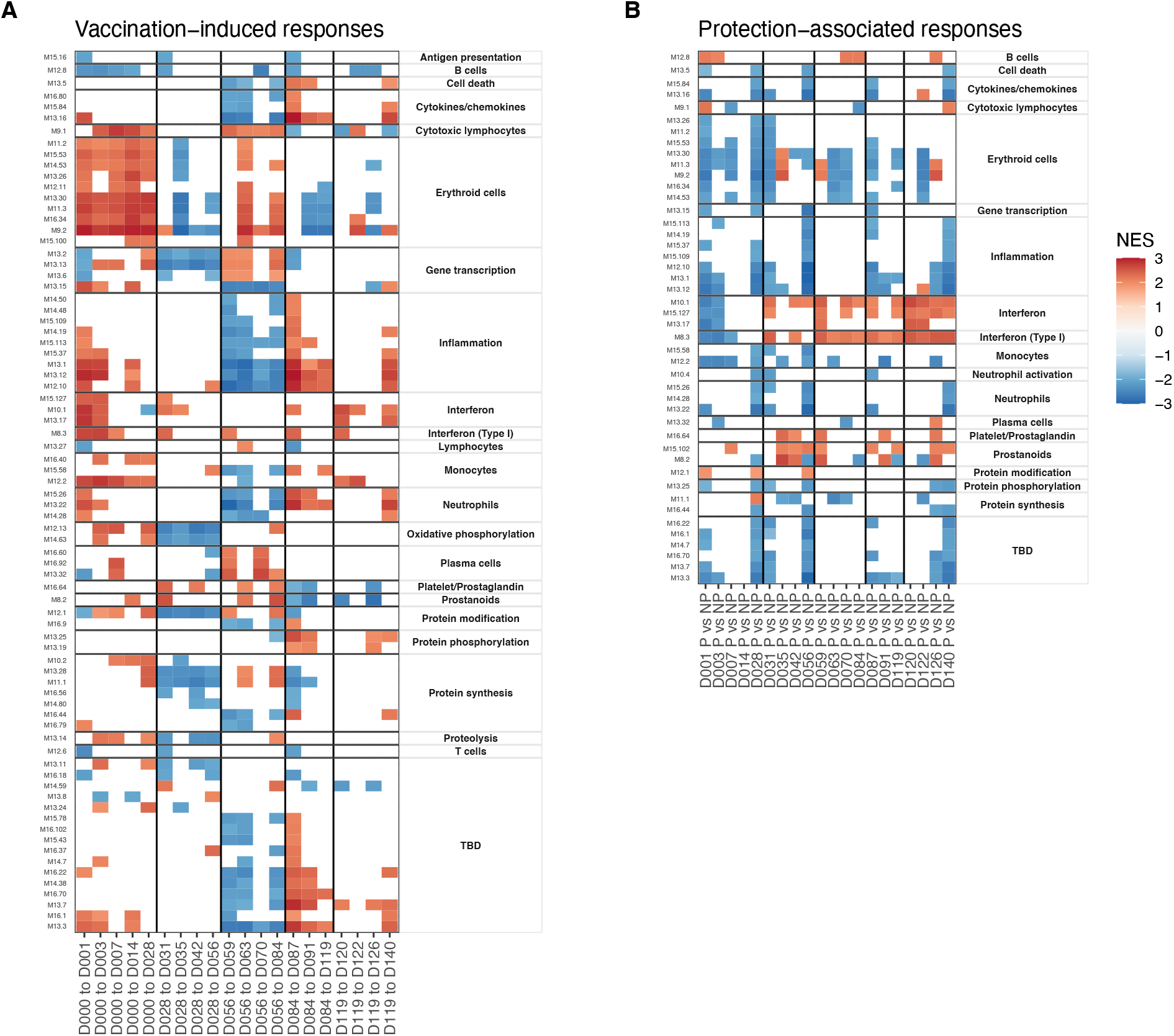
Gene set enrichment analyses showing patterns of vaccine-induced and protection-associated responses profiled using blood transcription modules (BTMs). Heatmaps show significant (FDR < 0.05) normalized enrichment scores (NES). **A**. BTMs significantly enriched for genes changing in expression relative to day of the most recent immunization. **B**. BTMs significantly enriched for genes with expression differences between protected and non-protected individuals at each timepoint. Vertical black lines indicate times of immunizations in both panels

Focusing on protection-associated BTMs, we saw more consistent patterns after each immunization. The majority of protection-associated processes comprised BTMs annotated as associated with specific immune cell types (adaptive and innate immune cells), cytokines, erythroid cell and inflammation and proliferation (protein synthesis/phosphorylation/modification) that were consistently reduced in protected individuals relative to non-protected individuals 1-3 days after each immunization (**Figure 3 B**). As with the vaccine-responsive BTMs above, ‘TBD’ modules mimicked the protection-associated enrichment patterns of inflammation associated BTMs. Interferon and prostanoid BTMs which were increased in protected participants immediately after immunizations 2-5, but not the first immunization. These interferon responses are consistent with what was observed in the DEG analysis above.

### Hierarchical clustering revealed distinct patterns of protection-associated responses

In order to decipher the different patterns of protection-associated responses, we calculated average BTM expression and hierarchically clustered the modules based on their correlation matrix. BTMs that shared a functional annotation tended to cluster closely (**Figure 4 A**) with Interferon BTMs and erythroid cell BTMs forming highly distinct clusters. We also observed a distinct cluster that represented inflammation/neutrophil/cytokine BTMs. This indicated that BTMs representing similar functional annotations shared coherent expression patterns. Representative BTMs were then selected from these sub-clusters and their averaged expression was plotted (**Figure 4 B**). Alongside the previously noted type I Interferon BTM, we observed a variety of protection-associated expression patterns, including both transient responses to vaccination and sustained protection-associated differences over the study. Notably, M12.2 monocyte BTM expression showed strong increases in protected individuals shortly after immunizations 2-5. In contrast, the M11.3 erythroid cells and M8.2 prostanoid BTMs showed sustained increased expression in non-protected individuals throughout the study. Thus, GSEA revealed numerous functional gene sets and expression dynamics that were correlated with protection.

**Figure 4:**
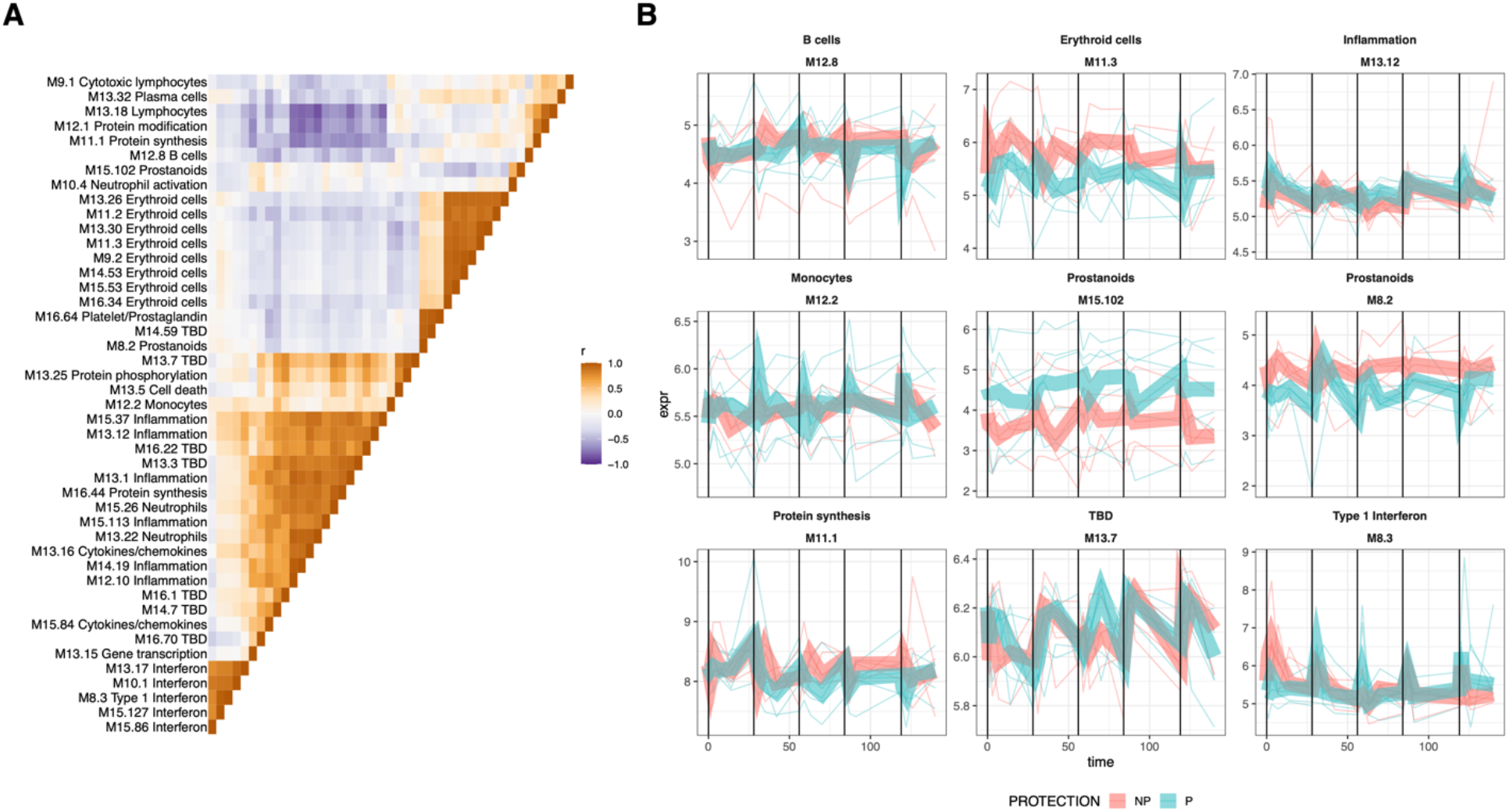
Expression dynamics of protection-associated BTMs. Expression of protection-associated BTMs identified by GSEA was calculated as the average of BTM gene expression and used to create a BTM correlation matrix. **A**. The BTM correlation matrix, with BTM-BTM correlation coefficients (Pearson’s r) indicated, and BTMs ordered identically on the x- and y-axes by hierarchical clustering of the correlation matrix. **B**. Averaged expression of representative BTMs from subclusters in **A**. plotted with lines representing individuals BTM expression, colored by protection status. Protection group average expression shown as thick ribbons

### Monocyte, Dendritic cell and invariant T cell populations were responsive to RAS vaccination

To better understand the cellular RAS vaccination response profile, we performed high-parameter flow cytometry profiling of PBMCs taken from the IMRAS vaccinees at timepoints overlapping with whole blood gene expression (**Table 1**). Four protein marker panels were used, consisting of 21-28 markers each, specific for B cells, T cells, antigen-presenting cells, and NK/ILC/ γδT T cells (**Table S 1**). An extensive gating strategy was applied (see Methods) to determine counts for hundreds of different cell types and corresponding activation states (**Datafile S 2**). As with whole-blood transcriptomics, cell abundance profile variability was most strongly predicted by person-to-person variability (**Figure S 2**) rather than by vaccine-induced changes, which necessitated a mixed-modelling approach to account for this variability while identifying vaccine-induced cell subsets.

Mixed-modelling revealed vaccine-induced cell types (**Figure 5 A, Datafile S 3**) including Vδ2 γδ T (**Figure 5 B**) cell subsets, classical CD14+ monocyte subsets (**Figure 5 C**), non-classical (NC) monocyte subsets (CD11c+ HLA-DR+ CD16+) (**Figure 5 D**) MAIT cell subsets, (**Figure 5 E**), and CD4+ and CD8+ T cell subsets. We observed few or no changes in B cell, NK cell or NKT cell subsets. Several cell type groupings showed coherent changes in abundance over the course of vaccination: NC monocyte subsets were rapidly and transiently increased after immunizations 1 and 3 and 5 in both protected and non-protected individuals. Larger increases were apparent in non-protected individuals after the first immunization with smaller increases after the third immunization, while the opposite pattern was observed in protected individuals (**Figure 5 B**). In contrast, Vδ2 γδ T cell subsets continuously increased over the course of vaccination and this increase was more apparent in non-protected individuals (**Figure 5 C**). Similar to NC monocytes, classical CD14+ monocytes subsets showed transient increases after each immunization (**Figure 5 D**). CD272- (a.k.a. BTLA: B-and T-lymphocyte attenuator) and CD85k+ (a.k.a. LILRB4, leukocyte immunoglobulin-like receptor B4) CD14+ monocytes represented the most abundant induced monocyte populations. For both CD272- and CD85k+ monocytes, increased abundance was apparent at the earliest post immunization 1 timepoint (day 3) in non-protected individuals, but not until day 7 post-immunization for protected individuals. A distinct pattern was observed for MAIT cell abundance. Total MAIT cell abundance was generally increased after each immunization for both protected and non-protected individuals. However, increases in total and CD8+ MAIT cells were stronger and more rapid in protected individuals after the first immunization. (**Figure 5 E**). Vaccine-induced T cell subsets, both CD4+ and CD8+, were not confined to a single memory or effector phenotype. However, since malaria sporozoite-specific T cells represent a small (or zero) proportion of total T cells it is unclear what is driving changes in circulating T cell proportion. These changes may simply reflect changes in the total number of circulating myeloid cells in response to vaccination.

**Figure 5:**
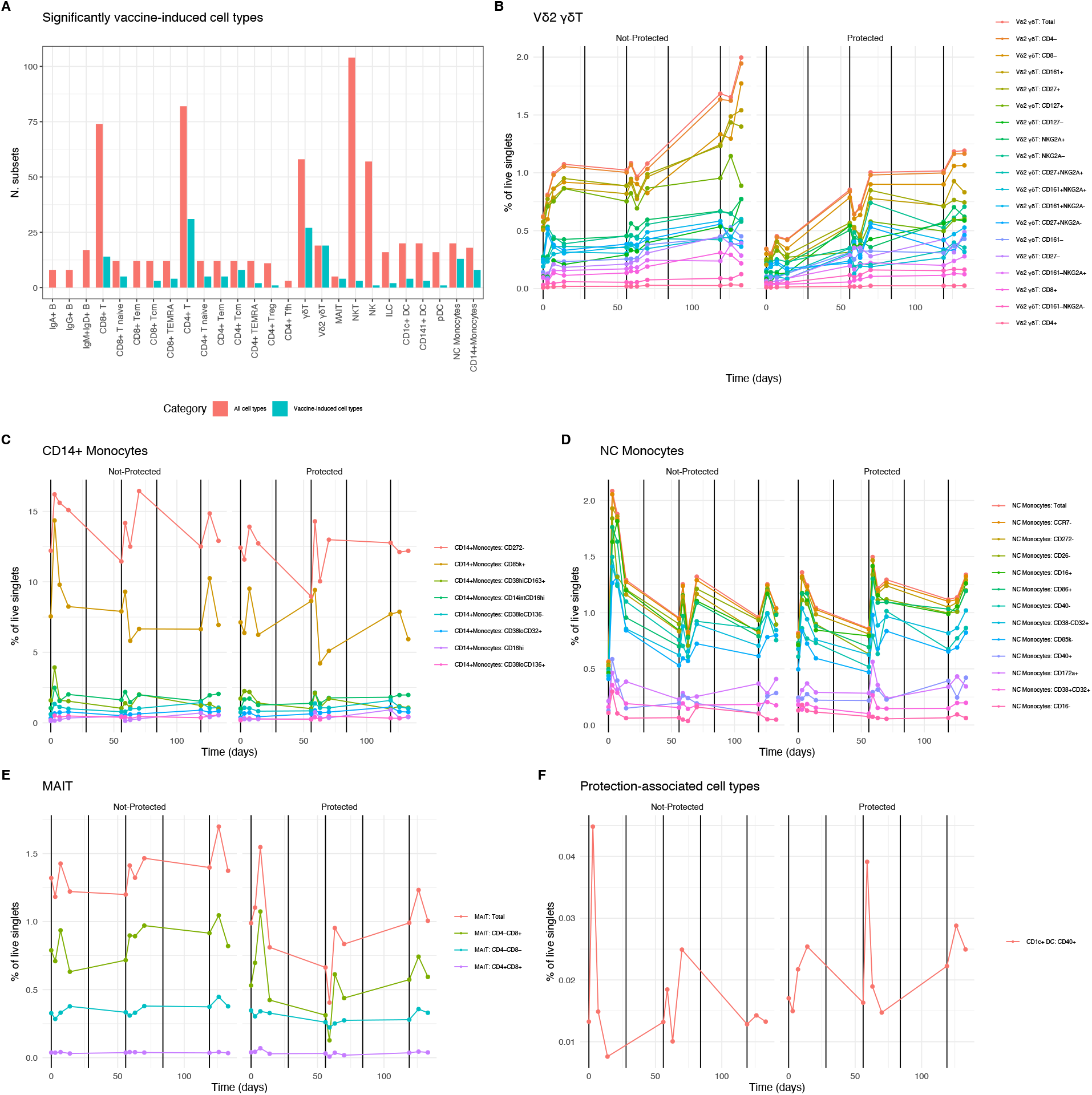
Cell type responses to RAS vaccination. **A**. Counts of significantly (FDR < 0.05) vaccine-induced cell types grouped by lineage and/or function. Red columns indicate total numbers of gated subsets in the corresponding group, adjacent teal columns show proportion of gated subsets that were vaccine-induced. **B-E** Averaged counts of vaccine-induced cell types showing vaccine induced cell types from specific lineage/function groupings in **A**., over time. Vertical black lines indicate vaccination times. Legend shows color for each gated cell type subset, ordered from most abundant (top) to least abundant (bottom). **B**. Vδ2 γδT cells, **C**. Classical CD14+ monocytes, **D**. Non-classical (NC) monocytes, **E**. MAIT cells. **F**. Averaged counts of the protection-associated (FDR < 0.05) cell type: CD1c+ CD40+ DCs.

It is important to note that none of the protection-associated trends listed in the vaccine-induced cell types described above reached statistical significance. We identified a single significantly protection-associated cell type: CD40+ CD1c+ DCs (**Figure 5 F**). These showed a similar overall expression pattern as the NC monocytes, but with a more pronounced increase in abundance in non-protected individuals after the first immunization and a more pronounced spike in protected individuals after the third immunization. In summary, we observed that myeloid cell types including classical CD14+ monocytes, non-classical monocytes and DCs tended to show similar vaccine response and protection dynamics as type I interferon-associated gene expression, while other vaccine responsive lymphoid cell types such as MAITs and Vδ2 γδT cells had distinct response patterns over time.

### Combined analysis of cell populations and gene expression revealed cell types correlated with vaccine-induced and protection-associated transcriptional responses

To better understand the relationships between cell type abundances and transcriptional responses after RAS immunization, we created a comprehensive correlation map relating RAS induced BTM expression to cell type abundances. Correlations between BTM expression and vaccine-induced cell type abundances were calculated by matching transcriptomic profiles with flow-cytometry derived cell type abundanced from each participant at shared timepoints (**Table 1**). Multiple significant cell type-BTM correlated pairs were observed (**Figure 6 A**), and these correlations often reflected known functions of the cell type in question. For example, BTMs with the ‘inflammation’ functional annotation were correlated with CD14+ monocyte subsets and BTMs with the ‘cytotoxic lymphocyte’ annotation were correlated with γδ T cells. Many BTM functionally annotated as ‘antigen presentation’, ‘lymphocytes’; ‘T cells’, and ‘cell cycle’ were specifically correlated with naïve CD4+ and CD8+ T cell subset abundance (**Figure 6 B**), possibly reflecting initiation of adaptive responses post immunization.

**Figure 6:**
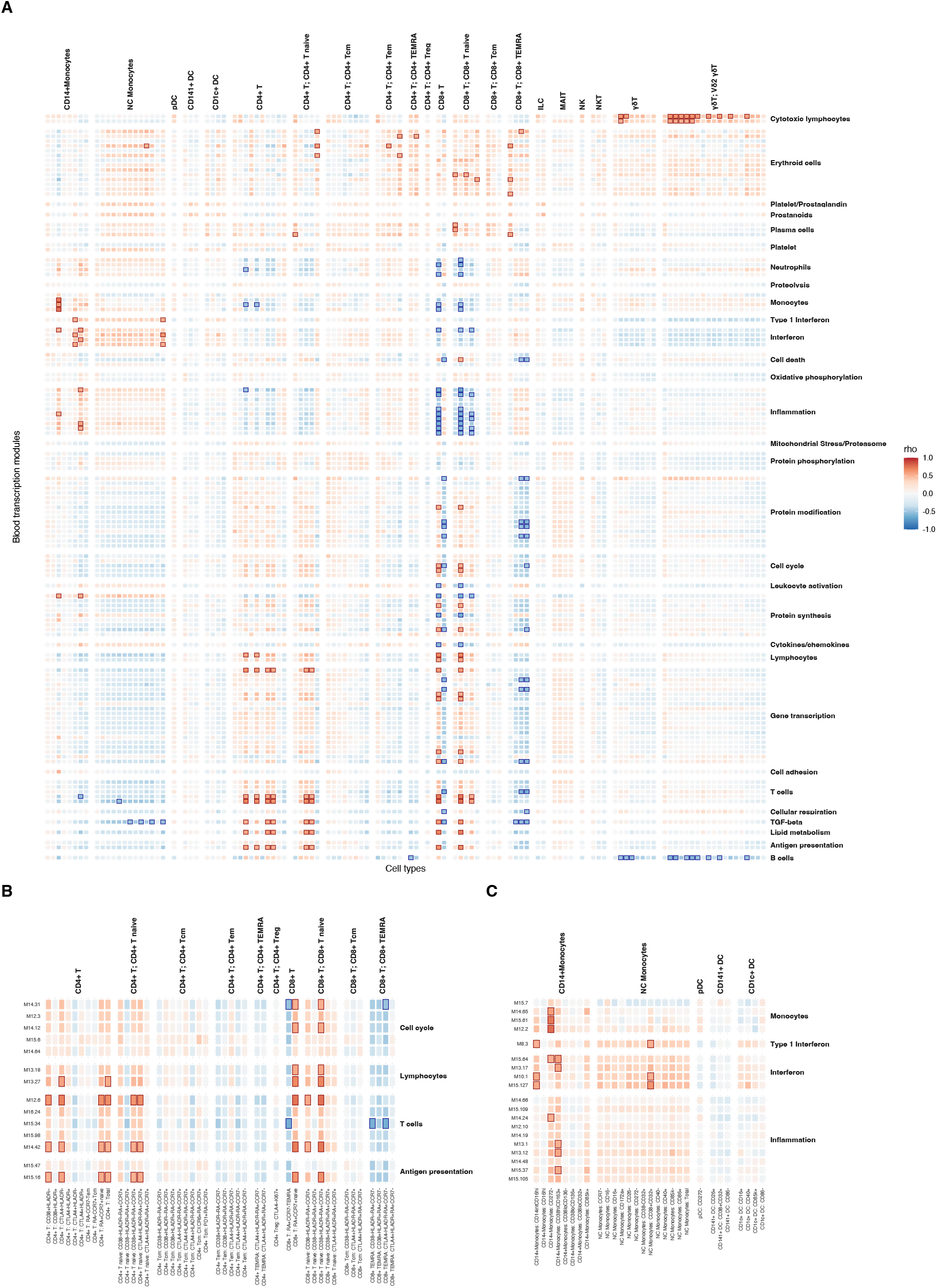
Correlations between vaccine-induced cell type abundances and vaccine-induced BTM expression. **A**. Heatmap showing Spearman correlations between vaccine-induced cell type abundance and average BTM expression. Only BTMs with at least one significant cell type correlation are shown. Strong positive and negative (|ρ| > 0.5 & FDR < 0.05) correlations are highlighted with red and blue boxes respectively. **B**. Subset of heatmap **A**. with T cell subsets labelled. **C**. Subset of heatmap **A**. with DC and Monocyte subsets labelled.

Several monocyte subsets were significantly (FDR < 0.05) and strongly positively correlated (Spearman’s ρ > 0.5) with Interferon BTM expression, specifically the CD16hi, CD272- and CD38hiCD163+ subsets of CD14+ monocytes, and CD38+CD32+ NC monocytes (**Figure 6 C**). Vaccine-induced NC monocyte subsets were uniformly positively correlated (all NC monocyte – interferon ρ > 0.2) with all 4 interferon BTMs. While most CD14+ monocyte populations were positively correlated with interferon BTMs, this was not always the case for CD38lo CD32+ or CD16hi populations of CD14+ monocytes. Therefore, it appeared that NC monocyte abundance was linked to interferon expression in the blood across the entire study period regardless of NC monocyte activation/maturity status, while this was not the case for CD14+ monocyte populations.

To further explore the temporal linkage between interferon transcriptional responses and DC/monocyte abundance, we investigated how the strength of their correlation varied over time. After immunization 1 (day 0-28) specifically, 2 CD14+ monocyte subsets were significantly positively correlated with interferon BTM expression. In contrast only 1 NC monocyte subset and no CD1c+ DCs (**Table 2**) were correlated with interferon BTMs. This changed after immunization 3, when 9 NC monocyte subsets, including total NC monocyte counts, were significantly correlated with interferon along with 3 CD14+ monocyte subsets and one CD1c+ DC subset (the protection associated CD1c+ CD40+ DC subset). This suggests that non-protection associated interferon responses after immunization 1 are primarily associated with subsets of CD14+ monocytes, while protection-associated interferon responses after immunization 3 were correlated with increased abundance of a wider spectrum of both CD14+ and NC monocyte populations.

**Table 2:** Numbers of DC/monocyte subsets significantly (FDR < 0.05) and positively (rho>0.25) correlated with at least one interferon BTM after each immunization.

|  | All timepoints<br>(Day 0 -140) | Immunization 1<br>(Day 0 - 28) | Immunization 3<br>(Day 56 - 81) | Immunization 5<br>(Day 119-140) |
| --- | --- | --- | --- | --- |
| **CD14+Monocytes** | CD14intCD16hi<br>CD272-<br>CD38hiCD163+<br>CD85k+ | CD272-<br>CD38hiCD163+ | CD14intCD16hi<br>CD272-<br>CD38hiCD163+ |  |
| **CD1c+ DC** | CD16+<br>CD40+ |  | CD40+ |  |
| **NC Monocytes** | Total<br>CCR7-<br>CD16-<br>CD16+<br>CD26-<br>CD272-<br>CD38-CD32+<br>CD38+CD32+<br>CD40-<br>CD40+<br>CD85k-<br>CD86+ | CD38+CD32+ | Total<br>CCR7-<br>CD16+<br>CD26-<br>CD272-<br>CD38+CD32+<br>CD40+<br>CD85k-<br>CD86+ |  |

## DISCUSSION AND CONCLUSIONS

Radiation-attenuated sporozoite vaccination is known to induce sterilizing protection against malaria, and many studies have investigated its protective efficacy and immune consequences [13,14,36–39]. Our work stands out as the most comprehensive profiling to date of radiation-attenuated sporozoite immunization and challenge in malaria naïve individuals. Previously [13], we reported that increased abundance of type I interferon-associated genes in blood were associated with a lack of protection in malaria-naïve RAS vaccinees after the first immunization. Here we extend our analysis by showing that interferon responses are subsequently increased more strongly in protected relative to non-protected individuals after subsequent boosting immunization. We also identified several other independent transcriptional pathways associated with RAS-induced malaria protection. Furthermore, high levels of interferon-associated gene transcription in blood after the first immunization were correlated with a small number of specific classical CD14+ monocyte subpopulations, while peripheral interferon responses to subsequent immunizations were correlated with increased abundance of broader range of classical and non-classical monocytes, along with CD1c+ dendritic cells.

Serum antibody profiling revealed robust serum antibody responses to abundant sporozoite surface antigens, i.e. CSP, TRAP and AMA1, that were vaccine-induced but not protection-associated. We did not see significant changes in B cell subset abundances in blood induced by vaccination, or associated with populations, likely reflecting small total numbers of *P. falciparum* antigen specific B cells in circulation. The relationship between malaria protection and neutralizing antibodies is complex: despite a clear role for antibodies in protection against malaria, the association of serum antibody levels with protection is inconsistent [40,41]. It has been shown that malaria protection can be elicited by sufficiently high levels of neutralizing antibodies delivered via passive transfer [42], but It is likely that the antibody levels elicited in this IMRAS study cohort were insufficiently high to achieve protection alone, and require the participation of other mechanisms, such as malaria specific CD8+ T cells [43,44]. Also, our study is likely underpowered to discover different potential routes of protection from alternate antigens and to capture the breadth of antigen responses on an individual level. We did not explore Fc mediated antibody function, which has been previously associated with protection [40].

The biggest factor driving variability in transcriptome and circulating cell phenotype populations was individual variability. Each vaccinee occupied a distinct region of the dimensionality-reduced transcriptomic space (**Figures S 1, S 2**), illustrating that per-participant variability has a stronger effect than responses induced by vaccines. It is reasonable to assume that our limited sample size has not extensively explored common human transcriptomic phenotypes, and further insights into RAS immunity would be accessible by profiling larger cohorts. Nevertheless, significantly altered transcriptome responses and cell type abundances associated with vaccination and protection were evident. Vaccine-induced responses were highly distinct after each vaccination. We identified multiple coherently expressed transcriptional responses significantly responsive to immunization. Responses associated with vaccination, but not protection, primarily comprised strong reductions in proliferative processes (protein synthesis, gene transcription, protein phosphorylation) and oxidative phosphorylation BTMs after immunization 2. Metabolic processes in whole blood reflect the sum of each individual cell types’ metabolic program. As immune cells comprise the vast majority of nucleated blood cells, changes in metabolic gene expression in blood reflect changes in immune cell metabolism. Overall, vaccination-associated processes seem to represent broad shifts in immune cell metabolism and abundance after vaccination. Immature neutrophils use oxidative phosphorylation (OXPHOS) for energy production, while mature neutrophils rely on glycolysis [45]. Additionally, classically activated M1 macrophages and DCs upregulate glycolysis in preference to OXPHOS [46]. Thus, the reduction in OXPHOS and proliferative signaling after immunization 2 may reflect the increased presence of mature innate cell phenotypes. Additionally, OXPHOS is associated with anti-inflammatory M2 macrophages, while pro-inflammatory M1 macrophages rely on glycolysis [47]. Therefore, this reduction in OXPHOS after immunization 2 but not immunization 1 may reflect a shift in the balance of M1 vs M2 macrophages in the skin/site of infection after the first immunization. Alternatively, changes in OXPHOS signaling could be driven by T cell activation, as quiescent naïve and memory T cells primarily rely on OXPHOS, however activation via TCR stimulation causes a shift to glycolysis [48]. The large numbers of vaccine-induced genes observed immediately post-immunization contrasts with a recent whole-sporozoite with chloroquine prophylaxis vaccine study (CPS) with a similar number of participants and protective efficacy [8]. This is likely driven by the much smaller dose of sporozoites delivered: in the CPS study subjects received 5-15 bites from infected mosquitos per immunization vs. approximately 200 in IMRAS. However, BTMs implicated in protection largely overlapped between studies, including monocytes, neutrophils, inflammation and interferon.

Professional antigen presenting cells have an essential role in the development of vaccine-induced immunity, thus it is not surprising that monocyte and DC abundances were clearly induced by vaccination. In particular, multiple NC monocyte [30] subsets were increased after vaccination. In the original OMIP publication of the flow panel, these NC monocytes are referred to as double negative (DN) DCs, i.e. CD141- CD1c- CD14- CD11c+ cells which do not fall into the classical cDC1 or cDC2 classes and are not pDCs. However, we observed that almost all of these ‘DN DCs’ are CD16+. Previous work has shown that CD14dim CD16+ CD11c+ CD1c-CD141- cells are better described as non-classical monocytes and not DN DCs [49,50]. Thus, our results have consistently referred to non-classical (NC) vs. classical CD14+ monocytes. We saw increased abundance of classical CD14+ monocytes after the first immunization followed by increases in a broad mix of classical and non-classical monocytes after immunizations 3 and 5. Along with myeloid cells, RAS vaccination was associated with changes in non-conventional T cell populations. We saw increases in MAIT cell abundances in response to RAS immunization with a tendency towards larger increases in protected individuals. Mpina, et al. [51] have shown a long-lived, dose-dependent increase in MAIT cells following resolution of infection after CHMI with *ex-vivo* experiments that associated MAIT activation with blood stage malaria parasitemia, but not RAS administered by direct venous injection. It is therefore possible that MAIT cell responses detected here reflect immune responses to the mosquito bite, however this would not account for a trend towards larger increases in protected individuals, suggesting further investigation of the role of MAIT cells in malaria protection. Vδ2 γδT cells have been previously shown to be intrinsically activated by malaria [52,53] and elevated in protected individuals after whole-sporozoite vaccination[54]. We observed a consistent increase in Vδ2 γδT cell counts over the entire course of vaccination and non-protected individuals trended towards larger cumulative increases in these cells. This was particularly apparent after the fifth immunization, although this difference was not statistically significant. While this may appear at odds with the described protective role of Vδ2 γδT cells, other work suggests a complex relationship between Vδ2 γδT and malaria immunity: it has been reported that a decrease in Vδ2 γδT cells levels is associated with tolerance [55] and reduced severity of malaria infection[56].

We previously observed a detrimental role of type I interferon in the development of malaria protection after prime immunization and a correlation between it and non-classical monocytes. In the current study, we have further explored the dynamics of interferon response following RAS vaccination and observed that type I interferon was negatively associated with the initial establishment of adaptive memory, but positively correlated with memory expansion upon boosting. We also saw that these interferon responses were significantly correlated with monocyte frequency, consistent with our previous findings. A potential explanation for this phenomenon is that over-induction of type I interferon inhibits the initial establishment of memory cell populations. Sporozoite antigens are taken up and presented by DCs after caspase-mediated cell death [57], and type I interferon can inhibit caspase activity [58] and hence antigen presentation. The detrimental role of interferon in the development of CD8+ T cells mediated immunity has also been observed in animal models [44]. It remains to be seen whether the role of interferon in is specific to RAS immunization or is relevant to the development of human liver-stage malaria immunity more broadly. Radiation-attenuated sporozoites cannot replicate in the liver resulting in incomplete liver stage development without expansion of parasite mass. This is different in key respects to other whole-sporozoite vaccination approaches such as vaccination using late-arresting genetically attenuated sporozoites [11] or administration of live sporozoites with anti-malarial prophylaxis [8,12], which allow liver stage parasite expansion. Thus, the arrested nature of the RAS liver stage may explain the transient nature of the IFN transcriptional responses that peaked 1-3 days post immunization.

While abundances of CD14+ and NC monocyte populations were not significantly associated with protection, different monocyte subsets were correlated with interferon transcriptional responses after immunizations 1, 3 and 5, suggesting that the distinct protection-associated roles of interferon after immunization 1 vs immunizations 2-5 may reflect the balance of classical vs. non-classical monocytes responding to vaccination. Specifically, protection-associated interferon transcriptional responses were correlated with increased levels of both CD14+ and NC monocyte subsets, while the non-protective interferon responses after the 1^st^ immunization were correlated with CD14+ subsets only. Interestingly, increased abundance of non-classical CD14dim CD16+ monocytes has been previously linked to reduced serum inflammatory cytokine levels and less severe malaria in children [59], pointing to a key role for these monocyte subsets in resolving the inflammatory response to malaria. Thus, NC monocytes may play an immune regulatory role in response to RAS immunization, and effective vaccine mediated immunity may require a balance between classical and non-classical monocyte responses. The sole protection-associated cell type we identified was CD40+ CD1c+ DCs, which are activated cDC2 cells. cDC2s are the predominant DC population in blood, and can cross present to and potently activate Th1, Th2, Th17 and CD8+ T cells [60]. CD40 is a co-stimulatory molecule that plays a central role in T and B cell activation and regulation when expressed on DCs [61]. CD40 has been shown to be required for liver-stage protective immunity to malaria infection in a mouse model [62]. It has also been observed that blood derived DCs upregulated CD40 upon exposure to P. falciparum infected red blood cells [63]. CD40+ CD1c+ DC abundances were also correlated with interferon responses across all timepoints and specifically after immunization 3 in pattern similar to many NC monocyte subsets, suggesting that NC monocytes and DCs may play a coordinated role mediating expansion of adaptive memory responses.

Apart from interferon-associated signaling, BTMs associated with inflammation, cytokines and chemokines, monocytes, neutrophils, and erythroid cells were identified as protection associated by GSEA. In general, protected individuals showed reduced induction of these processes immediately after every immunization, distinct from what we saw with interferon. However, it is difficult to disentangle the specific role each of these individual processes play in protection. Interestingly, we have seen that increased pre-vaccination abundance of BTMs associated with inflammation, monocytes, and neutrophils was associated with protection against malaria challenge [64], raising the possibility that the reduced responses seen in protected individuals post-immunization is due to their higher baseline expression of these modules, giving them an immunological ‘head start’ in responding to immunization more broadly. That hypothesis is supported by a recent cross-study analysis of 13 different vaccines showing that a heightened inflammatory state prior to immunization marked by increased expression of monocytes, dendritic cells and pro-inflammatory genes was associated with higher serum antibody responses across vaccines [65]. Multiple BTMs associated with erythroid cells were identified as increased and negatively associated with protection 1-3 days post-immunization after the first three immunizations. It is not immediately clear why erythroid cell-associated processes might be important for RAS-mediated protection, but there are several possible explanations. For example, increased erythroid cell BTMs may reflect anemia of inflammation [66] where inflammation induces upregulation of hepcidin in the liver, resulting in iron retention in macrophages and reduction in erythroid cell differentiation and proliferation via inhibition of erythropoietin. Also, it has been shown that type I interferon is involved in hematopoiesis [67,68] and activates dormant hematopoietic stem cells. Increased prostaglandin BTMs were also associated with protection and their role may intersect with erythroid cell signaling, as free heme impairs the anti-inflammatory prostaglandin E_2_ [69] while decreased levels of plasma-circulating prostaglandin E2 were correlated with more severe malaria disease in children [70].

Taken together, our systems immunology profiling results demonstrate that a finely tuned and coordinated set of immune responses are involved in establishing protection after RAS immunization, in particular the interplay between interferon transcriptional responses and classical and non-classical monocyte abundances. These also include responses previously reported, and observed here, to be induced by whole sporozoite vaccines (e.g. expansion of Vδ2 γδT cells) and more general inflammatory, antigen uptake and presentation responses necessary for development of immunity from any vaccine. This work has revealed that the coordination of protection-associated peripheral blood responses is complex, highly variable on an individual level and distinct after each vaccination. We hope that the unprecedented depth and resolution of our analyses will inform future design and interpretation of whole-sporozoite vaccine trials.

## Data Availability

Whole blood transcriptional data is available on NCBI GEO under accessions GSE116619 (Days 0-28) and GSE192756 (Days 31-140). Antibody microarray profiling results and flow cytometry subset counts are available as Datafiles S 4 and S 5 respectively, with raw data available in ImmPort.

## SUPPLEMENTARY MATERIAL

**Figure S 1.**
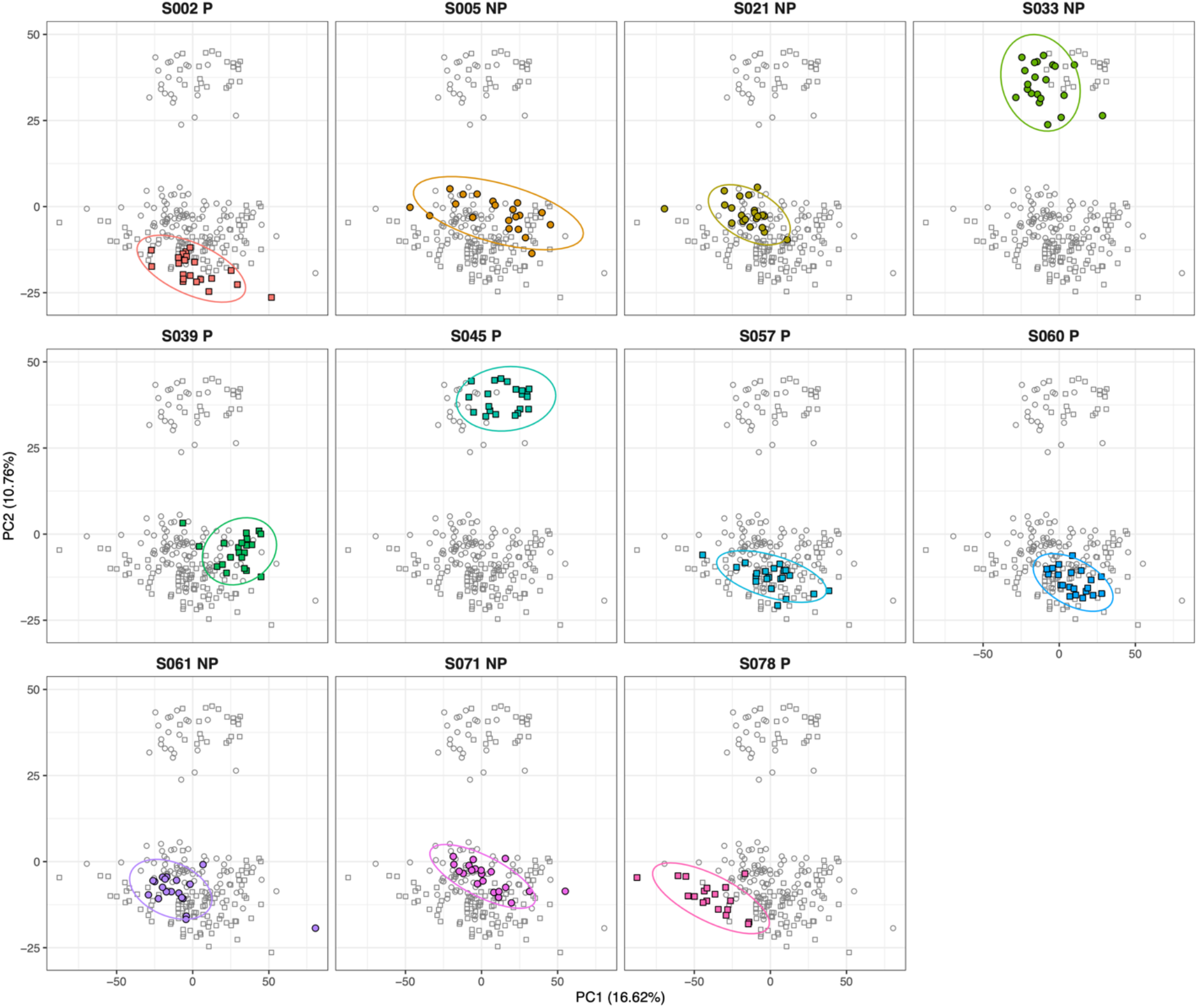
Principal component analysis plots of normalized whole blood gene expression data. Each panel shows colored points corresponding to all timepoints from a single participant with samples from other study participants shown in grey.

**Figure S 2:**
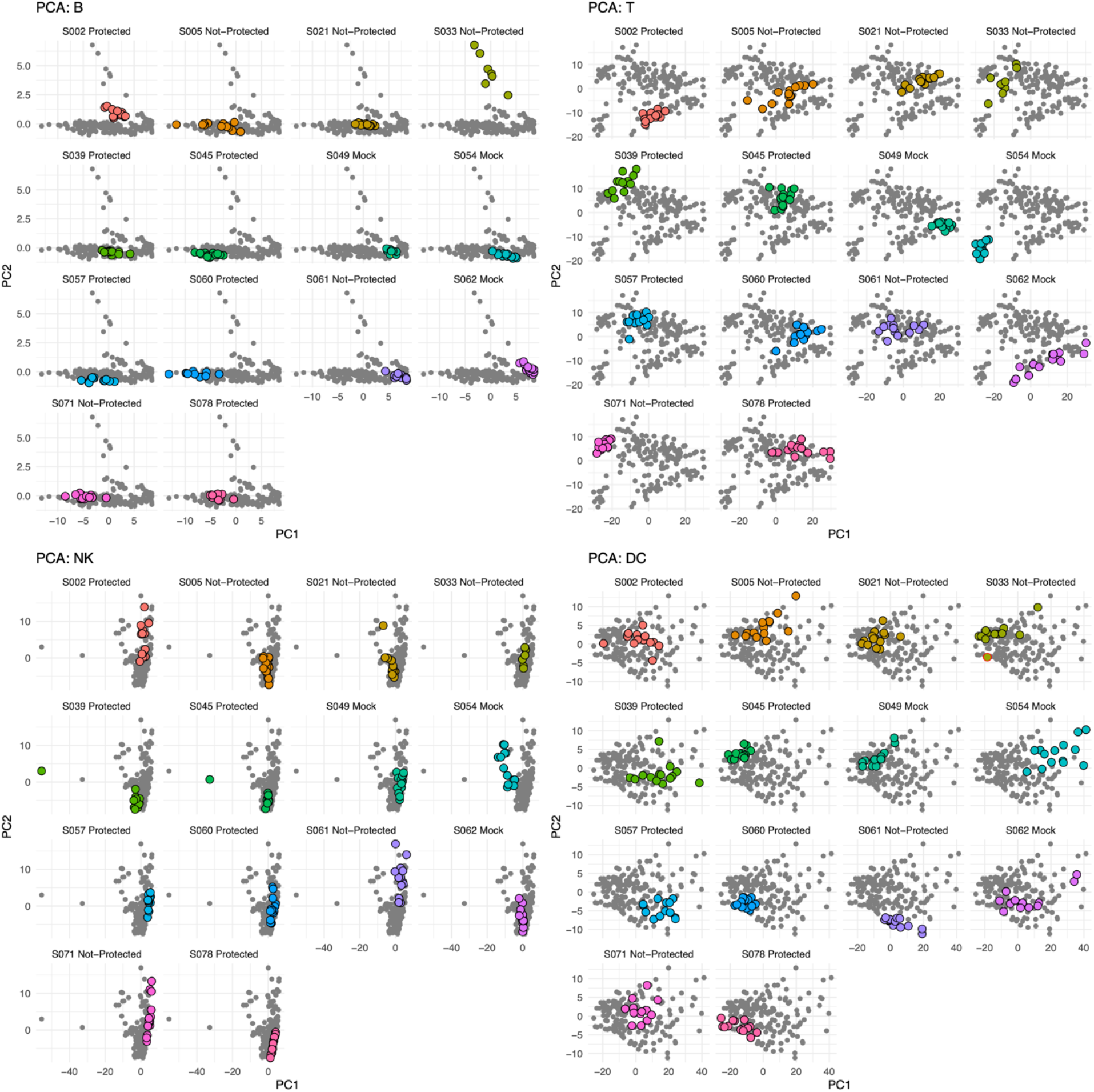
Principal component analysis plots of cell-type profiles determined by flow-cytometry. Each plot panel shows the first two principal components per-sample from flow panels **for** B-cells; CD3+ T cells; NK and invariant T-cells; **and** DCs and other antigen presenting cells respectively. Each sub-panel is labelled and colored to highlight a single study participant, with the remaining samples shown in grey.

**Table S1:**
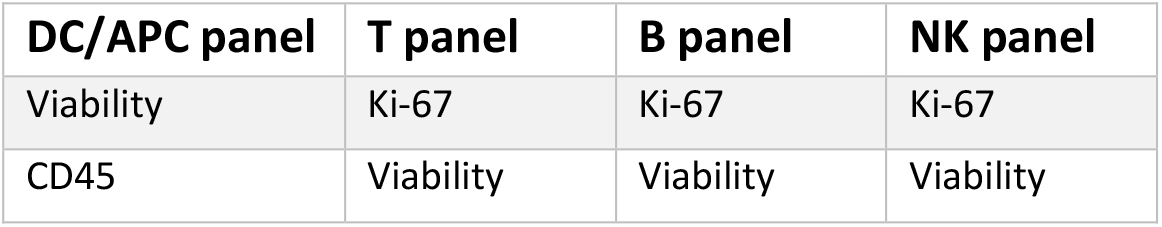
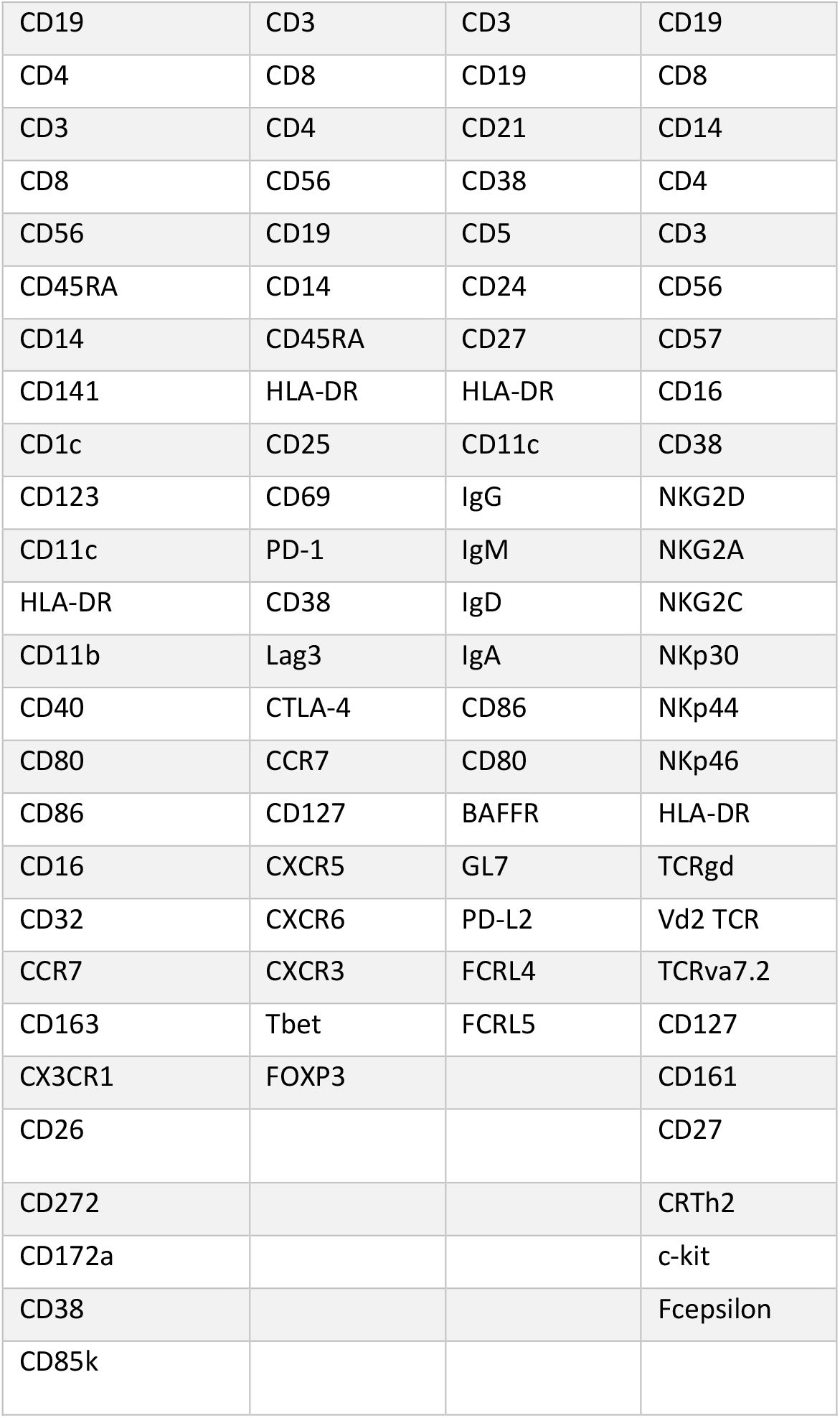
Flow cytometry panel marker panels. Each column lists the protein markers used in each flow-cytometry panel in this study

| DC/APC panel | T panel | B panel | NK panel |
| --- | --- | --- | --- |
| Viability | Ki-67 | Ki-67 | Ki-67 |
| CD45 | Viability | Viability | Viability |
| CD19 | CD3 | CD3 | CD19 |
| CD4 | CD8 | CD19 | CD8 |
| CD3 | CD4 | CD21 | CD14 |
| CD8 | CD56 | CD38 | CD4 |
| CD56 | CD19 | CD5 | CD3 |
| CD45RA | CD14 | CD24 | CD56 |
| CD14 | CD45RA | CD27 | CD57 |
| CD141 | HLA-DR | HLA-DR | CD16 |
| CD1c | CD25 | CD11c | CD38 |
| CD123 | CD69 | IgG | NKG2D |
| CD11c | PD-1 | IgM | NKG2A |
| HLA-DR | CD38 | IgD | NKG2C |
| CD11b | Lag3 | IgA | NKp30 |
| CD40 | CTLA-4 | CD86 | NKp44 |
| CD80 | CCR7 | CD80 | NKp46 |
| CD86 | CD127 | BAFFR | HLA-DR |
| CD16 | CXCR5 | GL7 | TCRgd |
| CD32 | CXCR6 | PD-L2 | Vd2 TCR |
| CCR7 | CXCR3 | FCRL4 | TCRva7.2 |
| CD163 | Tbet | FCRL5 | CD127 |
| CX3CR1 | FOXP3 |  | CD161 |
| CD26 |  |  | CD27 |
| CD272 |  |  | CRTh2 |
| CD172a |  |  | c-kit |
| CD38 |  |  | Fcepsilon |
| CD85k |  |  |  |

**Datafile S 1**: Limma-dream results table of significant (FDR < 0.05) vaccine-induced genes

**Datafile S 2**: Gating diagrams for each flow cytometry panel

**Datafile S 3**: Mixed-model p-values and high-level cell type grouping for each vaccine-induced cell type

**Datafile S 4**: Serum antibody array mean fluorescent intensity (MFI) array profiling data for each antigen and sample, normalized relative to IVTT controls.

**Datafile S 5**: Gated flow cytometry count table, annotated with cell types for each participant at each timepoint

## DATA AVAILABILITY

Whole blood transcriptional data is available on NCBI GEO under accessions GSE116619 (Days 0-28) and GSE192756 (Days 31-140). Antibody microarray profiling results and flow cytometry subset counts are available as **Datafiles S 4** and **S 5** respectively, with raw data available in ImmPort.

## ACKNOWLEDGEMENTS

We would like to acknowledge all the IMRAS study participants who made this work possible. We thank the Naval Medical Research Center (NMRC) Malaria Department, the Clinical Trials Center, and the Clinical Immunology Laboratory staff led by Dr. Eileen Villasante, Dr. Judith Epstein, Dr. Martha Sedegah, respectively, that conducted the IMRAS clinical trial and processed, cryopreserved, inventoried and transferred the clinical samples.

