## Supplementary figures and images for "Longitudinal immune profiling after radiation-attenuated sporozoite vaccination reveals coordinated immune processes correlated with malaria protection"

### Datafile-S-2.pdf

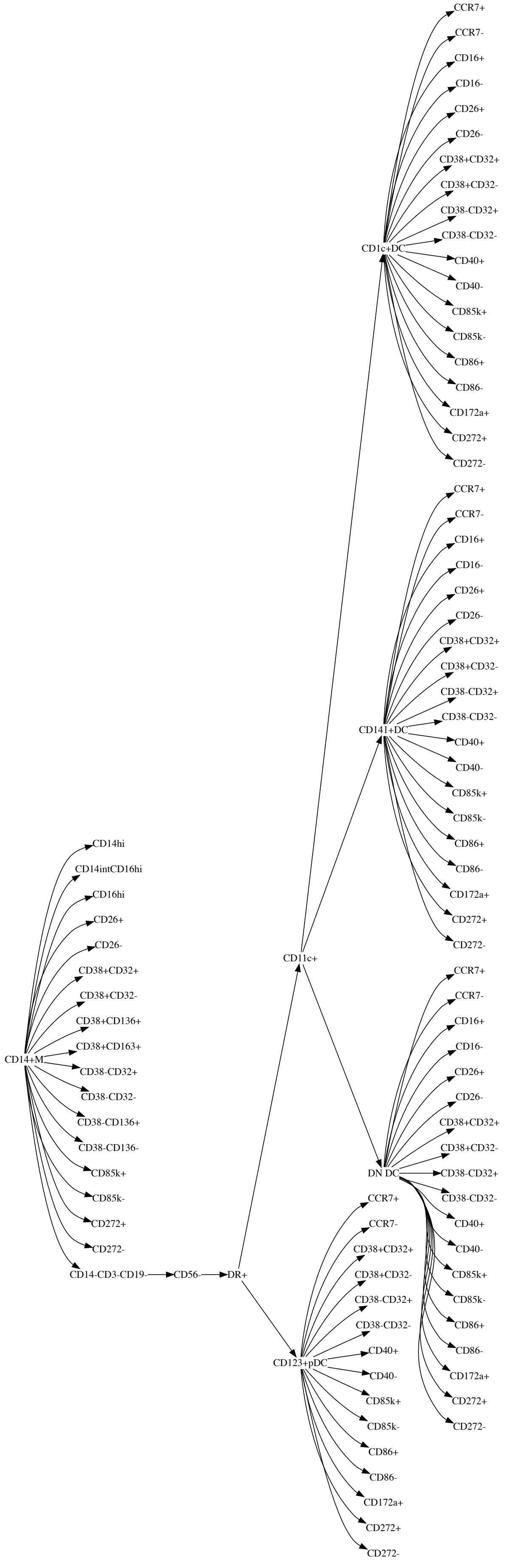


#### DC/APC gating

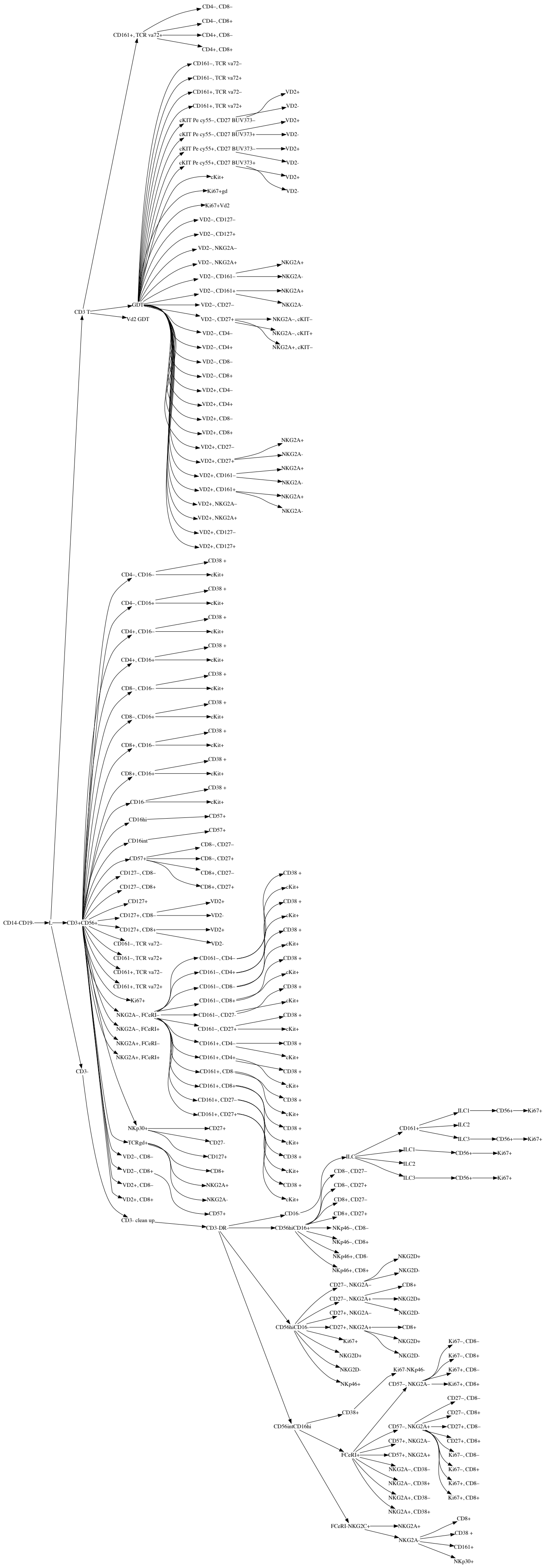


#### NK/ILC gating

T cell gating

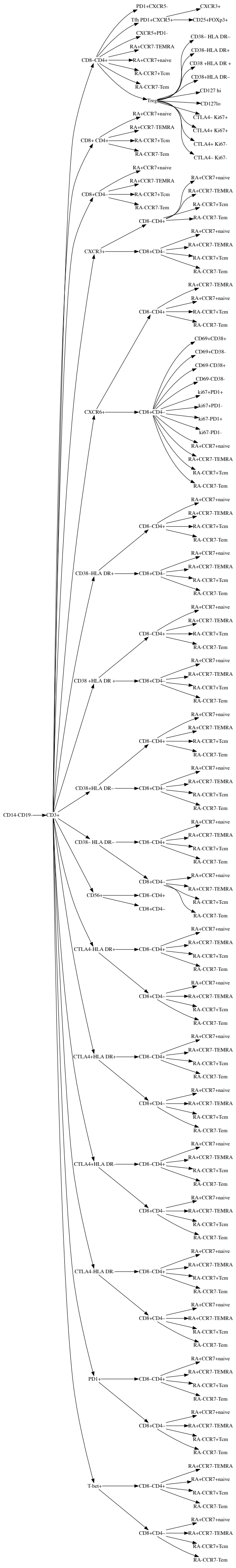

#### B cell gating

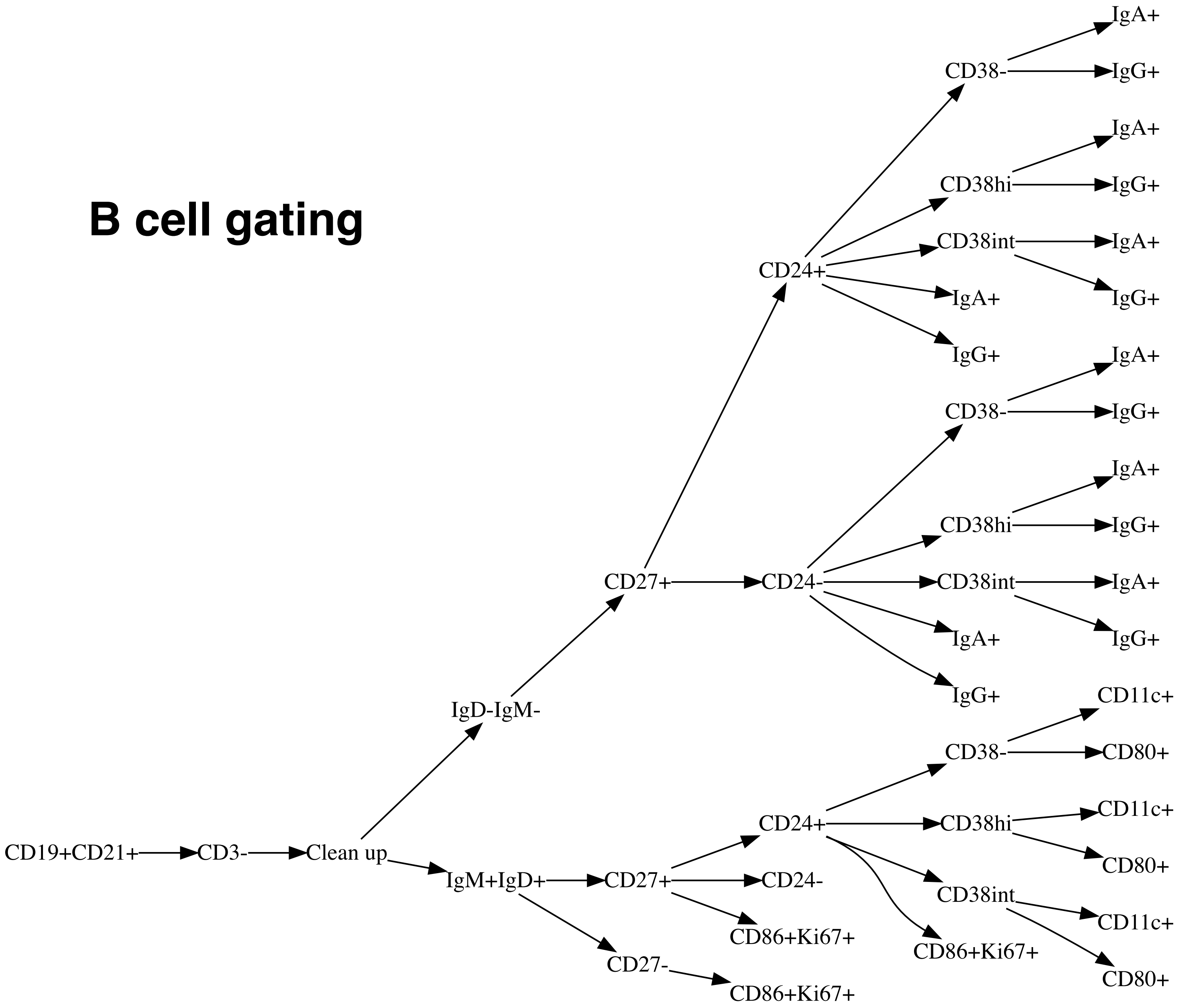
